# Estimation of heterogeneous instantaneous reproduction numbers with application to characterize SARS-CoV-2 transmission in Massachusetts counties

**DOI:** 10.1101/2021.12.02.21267164

**Authors:** Zhenwei Zhou, Eric Kolaczyk, Robin Thompson, Laura White

**Affiliations:** Department of Biostatistics, Boston University, Boston, Massachusetts, US; Department of Mathematics & Statistics, Boston University, Boston, Massachusetts, US; Mathematics Institute and SBIDER, University of Warwick, Coventry, England, UK

## Abstract

The reproductive number is an important metric that has been widely used to quantify the infectiousness of communicable diseases. The time-varying instantaneous reproductive number is useful for monitoring the real time dynamics of a disease to inform policy making for disease control. Local estimation of this metric, for instance at a county or city level, allows for more targeted interventions to curb transmission. However, simultaneous estimation of local reproductive numbers must account for potential sources of heterogeneity in these time-varying quantities – a key element of which is human mobility. We develop a statistical method that incorporates human mobility between multiple regions for estimating region-specific instantaneous reproductive numbers. The model also can account for exogenous cases imported from outside of the regions of interest. We propose two approaches to estimate the reproductive numbers, with mobility data used to adjust incidence in the first approach and to inform a formal priori distribution in the second (Bayesian) approach. Through a simulation study, we show that region-specific reproductive numbers can be well estimated if human mobility is reasonably well approximated by available data. We use this approach to estimate the instantaneous reproductive numbers of COVID-19 for 14 counties in Massachusetts using CDC case report data and the human mobility data collected by SafeGraph. We found that, accounting for mobility, our method produces estimates of reproductive numbers that are distinct across counties. In contrast, independent estimation of county-level reproductive numbers tends to produce similar values, as trends in county case-counts for the state are fairly concordant. These approaches can also be used to estimate any heterogeneity in transmission, for instance, age-dependent instantaneous reproductive number estimates. As people are more mobile and interact frequently in ways that permit transmission, it is important to account for this in the estimation of the reproductive number.

**Author summary:** To control the spreading of an infectious disease, it is very important to understand the real-time infectiousness of the pathogen that causes the disease. An existing metric called instantaneous reproductive number is often used to quantify the average number of secondary cases generated by individuals who are infectious at a certain time point, assuming no changes to current conditions. In practice, we might be interested in using the metric to describe the infectiousness in multiple regions. However, this is challenging when there are visitors traveling between these regions, since this could lead to a misclassification of where an individual is actually infected and create biased estimates for the instantaneous reproductive numbers. We developed a method that takes account of human mobility to estimate the instantaneous reproductive numbers for multiple regions simultaneously, which could reveal the heterogeneity of the metric. This method aims to provide helpful information on region-specific infectiousness for disease control measures that focus on the region with higher pathogen infectiousness. This approach is also applicable for estimating the reproductive number in the presence of other sources of heterogeneity, including by age.

## Introduction

In the aftermath of the pandemic caused by the SARS-CoV-1 virus, the idea of using surveillance data to estimate reproductive numbers was introduced and popularized by the seminal paper by Wallinga and Teunis [1]. Subsequent methods have been developed that are better suited to real time estimation, particularly the approach to estimate the instantaneous reproductive number introduced by Fraser [2] and implemented in the popular EpiEstim R package [3, 4]. These methods have promise to be useful in surveillance and monitoring an epidemic, but as the pandemic caused by SARS-CoV-2 has demonstrated, there are still needed improvements to these approaches. Principal issues include accounting for reporting delays in the data, underreporting of cases, and heterogeneity in transmission by geography and by demographic factors, such as age. For these methods to be truly useful in the ongoing COVID-19 pandemic and for future events, these issues must be addressed. Work is being done on the first two issues. For instance, Li et al. [5], Gunther et al. [6] and Martinez et al. [7] propose solutions to the timeliness problem. Pitzer et al. [8] demonstrate the impact of reporting issues and White et al. [9] have shown how estimates of *R*(*t*) can be corrected with information on the reporting fraction of diseases. In this paper, we propose a framework for addressing heterogeneity in transmission, specifically due to human mobility, though our methods can be more generally applied.

Studies have shown that there is transmission heterogeneity in COVID-19, as well as other infectious diseases, which lead to a disproportional impact of the disease on some groups. Multiple studies have found strong evidence of strong heterogeneity wherein a small number of individuals are responsible for the vast majority of cases [10–12]. Additionally, in this COVID-19 pandemic, Sy et al. [13] have shown how mobility, such as subway usage, in NYC lead to disproportionate case burden among those who are not maintaining physical distance. This implies it would be more efficient if we could account for the heterogeneity and focus control efforts on the populations with highest transmission probabilities [14].

Many factors could contribute to the heterogeneity of virus transmission, including important systematic factors such as lower social economic status (SES) that disadvantage certain groups and could lead to higher probability of disease transmission. These factors often cluster geographically. The impact of these factors on the virus transmission could be reflected on the reproductive numbers. Ideally we will be able to discover the heterogeneity by examining the differences of the reproductive numbers between different regions. However, due to human mobility, the heterogeneity of the reproductive numbers among different regions could be masked. This is because human mobility could distribute the infectees by certain infectors to different regions, leading to misclassification of the incidence of one region to another.

We propose a framework for accounting for heterogeneity in disease transmission when estimating the time-varying instantaneous reproductive number for each region. This could help monitor changes in transmission to guide public health measures, for example, implementing more stringent disease control measures for the region with higher virus transmission. Our framework requires data to inform the source or patterns of heterogeneity. We focus on human mobility data to estimate the reproductive number for multiple regions or population groups. However, we note that the framework is suitable for for understanding the effects of other important factors on the heterogeneity of the reproductive number, such as age.

We present an analytical framework with two approaches to estimate the dynamics of transmission heterogeneity. If we believe that the heterogeneity of the reproductive numbers can be recovered by accounting for population mixing due to human mobility, and we have confidence that the human mobility data represent the mixing of incidence, we suggest to use an efficient and straight forward approach that adjusts the incidence prior to estimation. If instead we want to adjust for importation of cases and more accurately quantify the uncertainty associated with the use of human mobility data with standard errors, we propose a more flexible and computationally intensive Bayesian approach that is more appropriate.

## Materials and methods

### Overview

We propose two approaches to estimate instantaneous reproductive numbers that incorporate human mobility data to account for heterogeneity. Both approaches are based on the framework of a system of renewal equations that bring human mobility into consideration. The difference between these two approaches is how the estimation handles potential uncertainty in the human mobility data. These methods can be applied to other types of heterogeneity, such as differential age-mixing where one might use information on contact patterns between age groups. Our first approach simplifies the problem by assuming that human mobility data accurately represents the mixing patterns and corresponding incidence misclassification without error. In this setting, we propose an approach that extends the framework developed by Fraser et al. [2] to estimate the heterogeneous instantaneous reproductive number by adjusting the observed incidences for multiple regions using the human mobility data. In reality, there is likely some randomness in human mobility and we would typically wish to quantify the uncertainty due to other factors that might drive the heterogeneity of instantaneous reproductive numbers. For this setting, we use a system of renewal equations that incorporates human mobility data and estimate instantaneous reproductive numbers under a hierarchical Bayesian framework. Both approaches are evaluated by simulations, and implemented to estimate instantaneous reproductive numbers for all counties in Massachusetts, USA, during the COVID-19 pandemic together with human mobility data from SafeGraph.

### Data

The COVID-19 incidence data is provided by CDC case report [15] and we use incidence from July 2020 to March 2021 because testing and case reporting became more frequent and regular starting in July 2020. We aggregate confirmed cases in Massachusetts by date and county. Human mobility data is obtained from the multiscale dynamic human mobility flow dataset constructed and maintained by Kang et al. [16], who computed, aggregated and inferred the daily and weekly dynamic origin-to-destination (O-D) flow at three geographic scales (census tract, county and state) analysing anonymous mobile phone users’ visits to various places provided by SafeGraph [17].

### Notation

Suppose that we want to estimate an instantaneous reproductive number, denoted as *R*(*t*), for *J* stratum, where the stratum can be geographical regions, age groups, communities, etc. Let *N*_*j*_(*t*), *t* = 1, …, *T* be the number of new cases reported at time *t* for region *j*, and *m*_*j*_(*t*) = *E*[*N*_*j*_(*t*)], where *t* = 1 is the first observation time and *T* is the last time with available data. The distribution of serial intervals is denoted as *ω*(*τ* |*θ*), where *τ* is the interval between the times of disease onset in an infector-infectee pair, and *θ* is the parameters of the distribution. There are several assumptions for both approaches that we propose:

1. Serial interval and reproductive number are statistically independent;
2. Reproductive number follows a Poisson distribution;
3. All infectors appear before those they infected;
4. Individuals mix homogeneously;
5. Closed population;
6. Complete case reporting;
7. The serial interval is the same as the reporting interval (i.e. the time between case report dates in an infector-infectee pair).

### Instantaneous reproductive number

The instantaneous reproductive number, originally developed by Fraser et al. [2], estimates the average number of secondary cases generated by individuals who are infectious at time *t* assuming no changes to current conditions. When using the instantaneous reproductive number, the expected incidence at time *t*, which is denoted as *m*(*t*), can be expressed as the following renewal equation:

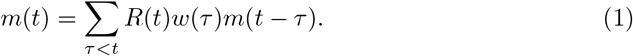

In practice, the estimator for *R*(*t*) can be computed with reported incidence *N* (*t*) as:

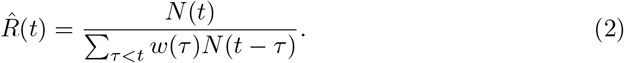

Cori et al. [3] use a Bayesian approach to estimate the *R*(*t*) with credible intervals and propose smoothing the estimates by using a longer time window, assuming the *R*(*t*) stay the same within that window. Thompson eto al. [4] extended the method to perform estimation in the presence of imported cases. Based on the renewal equation 1, and the estimation method developed by Cori et al. [3] and Thompson et al. [4] and with the assumption that the proportion of infected people are similar to mobility patterns of all individuals, we can formulate the process into a system of renewal equations that incorporates the human mobility data.

Denote *P* as the *J*-by-*J* human mobility matrix that reclassifies incidences to the presumed location of the transmission event. Let *p*_*j′j*_ be the entry of *P* matrix in the *j′*^*th*^ row and *j*^*th*^ column, and represents the proportion of population in *j*′ that travels to *j*. Then to describe incidences in multiple regions, we can extend the equation (1) to a system of equations:

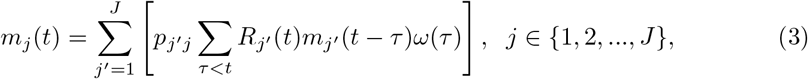

where 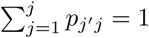. If we write it in a matrix form, we have:

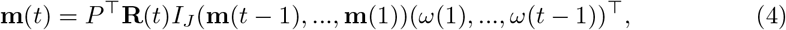

where **m**(*t*) = (*m*_1_(*t*), *m*_2_(*t*), …, *m*_*J*_ (*t*))^⊤^ is a vector of incidences for the *J* regions at time *t*, thus (**m**(*t* − 1), …, **m**(1)) is a *J*-by-(*t* − 1) matrix for the incidences of *J* regions from time *t* − 1 to 1. **R**(*t*) = (*R*_1_(*t*), *R*_2_(*t*), …, *R*_*J*_ (*t*))^⊤^ is a vector of instantaneous reproductive numbers for the *J* regions at time *t*.

Based on the above system of renewal equations, we propose two approaches for the estimation of heterogeneous *R*(*t*) incorporating mobility data as follows.

### Approach I – incidence adjustment approach

In this approach, we use the matrix *P* from the human mobility data deterministically. According to equation (4), assume that *P* is invertible, we have

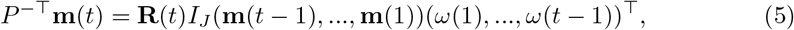

Note that

**m**(*t*) = (*m*_1_(*t*), *m*_2_(*t*), …, *m*_*J*_ (*t*))^⊤^ = (*E*[*N*_1_(*t*)], *E*[*N*_2_(*t*)], …, *E*[*N*_*J*_ (*t*)])^⊤^ = *E*[**N**(*t*)]. To estimate **R**(*t*) with the reported incidence **N**(*t*), let **N**_local_(*t*) = *P*^−⊤^**N**(*t*), and assume 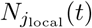 follows a Poisson distribution:

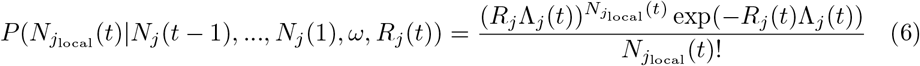

where 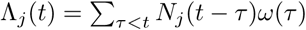

Assume that *R*_*j*_(*t*) follows a gamma prior distribution *Gamma*(*a, b*), and within a *k*-days window (from day *t* − *k* to *t*), the incidences all depend on the same *R*_*j*_(*t*). we can write the posterior of *R*_*j*_(*t*) as:

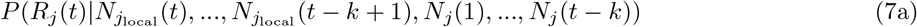

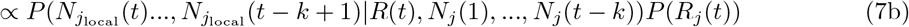

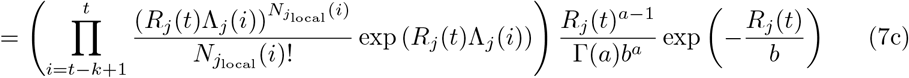

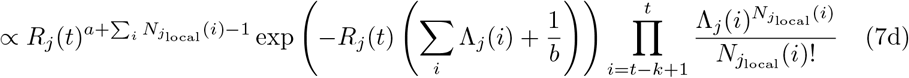

Thus, the posterior of *R*_*j*_(*t*) also follows a gamma distribution 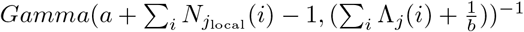. The estimation can be performed by mplementing the existing EpiEstim R package with the incidence adjustment data.

### Approach II – Bayesian approach

Based on the renewal equation with instantaneous reproductive number by previous studies [2] and [18], we formulate the renewal equations for *J* regions as:

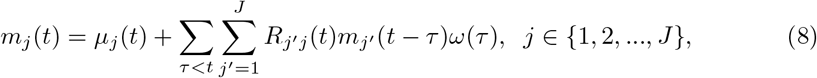

where *µ*_*j*_(*t*) is the rate of exogenous infections (infections out of any of the regions *j* ∈ {1, …, *J*}) occurs in region *j*, and *ω*(*τ*) is the probability distribution of serial interval. We model *R*_*j′j*_ (*t*) = *R*_*j′*_ (*t*)*p*_*j′j*_, where *R*_*j′*_ (*t*) is the region specific reproductive number for region *j*′ at time *t*, and *p*_*j′j*_ represents the probability of individuals in region *j* being infected by individuals in region *j*′, assuming that {*p*_*j′j*_ : *j*′, *j* ∈ 1, 2, …, *J*} are known. Then we have:

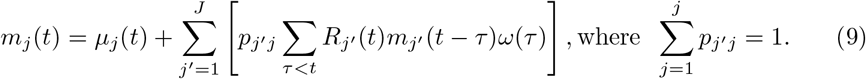

*p*_*j′j*_ here attempts to capture the information of transmission probability between the regions, and we denote a matrix *P* with entries *p*_*j′j*_ as a transition matrix that models the infected subjects flowing across the regions. For example, while estimating *R*(*t*) for multiple regions, we can inform the *P* matrix with mobility data between the regions and/or geographical distance between the regions. Within a Bayesian hierarchical modeling framework, Dirichlet priors for *P* can incorporate prior knowledge for the estimation of *R*(*t*).

We model *R*_*j′*_ (*t*) in (9) as:

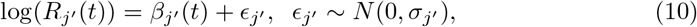

assuming *ϵ*_*j′*_ has constant variance over time.

Assume that the distribution of serial interval *ω*(*τ*) and *p*_*j′j*_ is known; *N*_*j*_(*t*) ∼ *Poisson*(*m*_*j*_(*t*)) and {*N*_*j*_(*t*)} are independent conditional on *m*_*j*_(*t*), so we have the factorization: 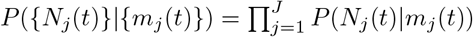. Then we can sample the posterior distribution of parameters with Bayesian hierarchical modeling:

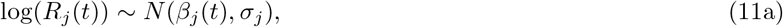

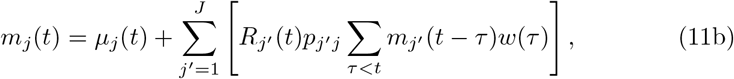

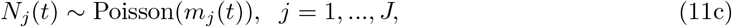

with certain prior specifications for {*µ*_*j*_ (*t*)}, {*β*_*j*_ (*t*)}, {*σ*_*j*_}.

We also allow a smoothing window for the estimation of *R*_*j*_(*t*). If the length of the smoothing window is *k*, then for time *t*_0_ we modify to be:

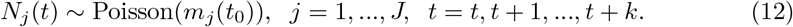

### Simulation

#### Simulation Settings

##### Scenario 1

We consider three regions (*j* ∈ {*a, b, c*}), where there are no exogenous infections (except for the initial cases on day 0), so that *µ*_*j*_(*t*) = 0. Assuming *p*_*j′j*_ and *ω*(*τ*) are known, we specify a 3-by-3 matrix *P* with entries *p*_*j′j*_, where *j*′ is row index and *j* is column index to approximate data we observe from [17]:

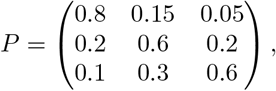

and we generate discrete distribution *ω*(*τ*) for *τ* from the CDF of *f* (*τ*) = *Gamma*(2, 0.5):

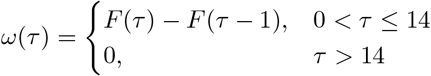

For {*R*_*j*_(*t*)}, we specify nonlinear functions for each region (also shown in Figure **??**):

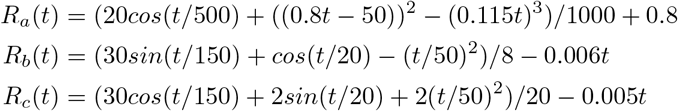

To generate incidence data, we let the initializing cases to be 10 in each region, and let the maximum time of observation to be *T* = 214. According to Equation 9, with {*R*_*j*_(*t*)}, *w*(*τ*) and matrix *P*, we can compute *m*_*j*_(*t*), then generate 100 replicates with *N*_*j*_(*t*) ∼ *Poisson*(*m*_*j*_(*t*)). Fig 1 shows the 100 Monte Carlo replicates of simulated data.

**Fig 1.**
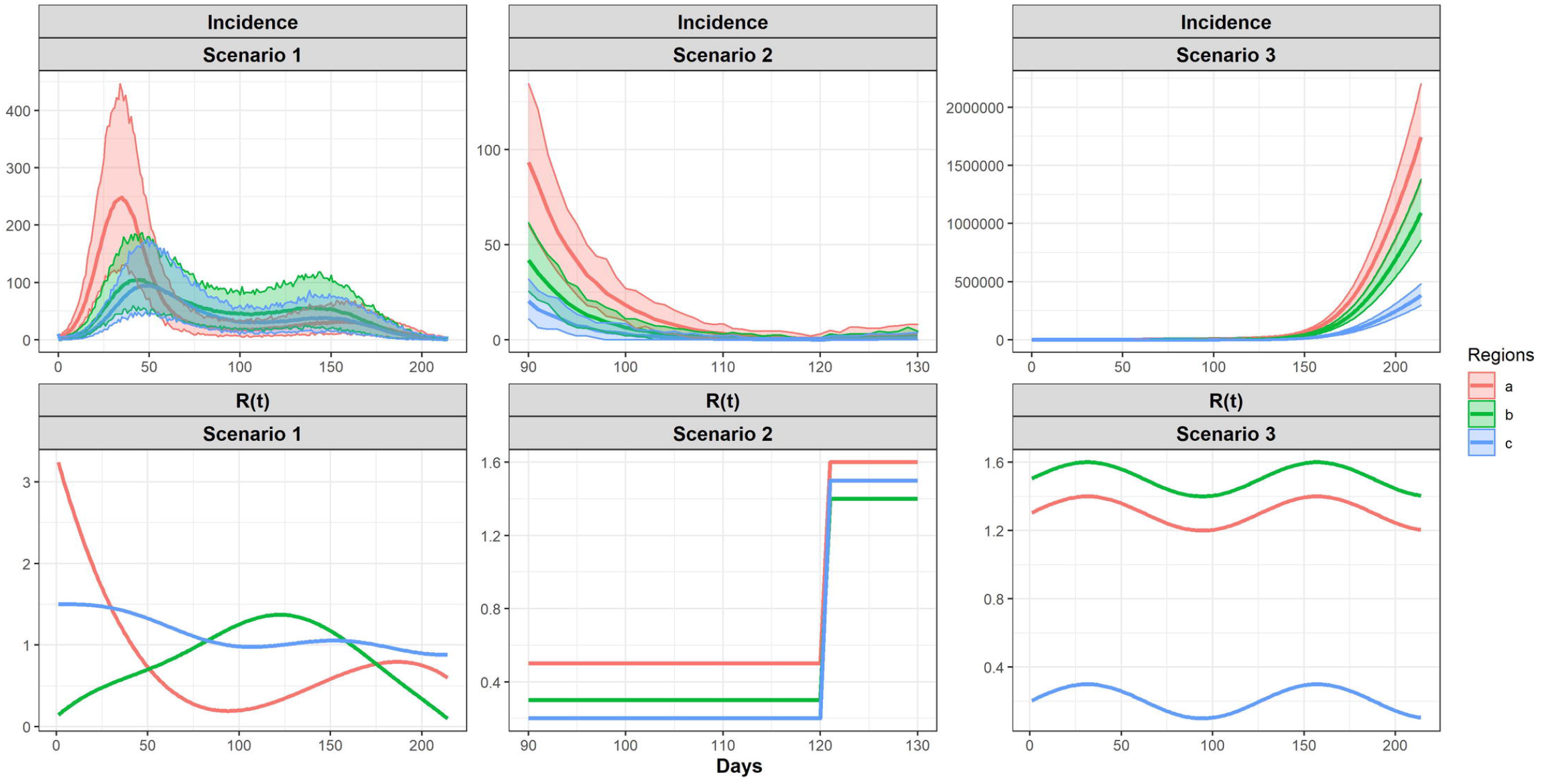
Specified *R*(*t*) functions and incidence for three regions from 100 replicates for simulation.

We performed the incidence adjustment approach (Approach I) to estimate the instantaneous reproductive numbers on the simulated data described above. For the Bayesian approach (Approach II), we evaluated the performance of the model using different distribution assumption for the incidence *N*_*j*_(*t*), and also using different lengths of smoothing window. Then we explored whether using a prior for the transition *P* matrix to allow for more flexibility could yield proper estimate for *R*_*j*_(*t*). The performance of the proposed model is compared with the model without considering the heterogeneous of *R*_*j*_(*t*), that is using an identity *P* matrix.

For all models, we use *N* (0, 0.5) as the prior distribution for *β*, and *N* (0, 1) for *σ*, other model parameter settings are described below:

Model 1: constant *P* matrix, smoothing window is 1, assume Poisson distribution for *N*_*j*_(*t*);

Model 2: constant *P* matrix, smoothing window is 8, assume Poisson distribution for *N*_*j*_(*t*);

Model 3: constant *P* matrix, smoothing window is 8, assume Negative Binomial distribution with *ϕ* ∼ *N* (0, 5) for *N*_*j*_(*t*);

Model 4: random *P* matrix that each column follows a Dirichlet distribution centering at the true *P* matrix with large concentration parameter, smoothing window is 8, assume Poisson distribution for *N*_*j*_(*t*);

Model 5: constant identity *P* matrix, smoothing window is 8, assume Poisson distribution for *N*_*j*_(*t*) (this model is equivalent to Fraser’s method, which do not consider human mobility);

The estimates from the Approach I (with and without mobility information) and model 4 and 5 of Approach II are shown in the main result section. Note that model 4 of Approach II is with mobility information, and model 5 of Approach II is without mobility information. Model 1, 2, 3 of Approach II are shown in Fig S1 in S1 Appendix.

##### Scenario 2

In practice, we might have a low count of cases for some of the regions, so we also evaluated the proposed approaches under the scenario where we have a lower count during certain period of time. In the low count scenario, we specify the *R*(*t*) for the three regions to be three piece-wise function, and it is shown in Fig 1.

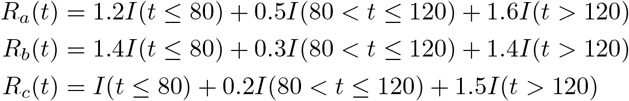

We initiate with 50 incidence for the tree regions, and simulated incidence from 100 replicates are shown in Fig 1. When performing estimation, we focus on the range from day 90 to day 130, because this region cover the period where the incidence decreases to zero or low counts that are close to zero. The result for estimated *R*(*t*) is shown in Fig S2 in S1 Appendix.

##### Scenario 3

We also evaluate the performance of the proposed approaches in a scenario where the population are traveling out of two of the regions with higher *R*(*t*) to the third region with lower *R*(*t*). We expect this scenario will show that if we do not consider the human mobility, we will over estimate the *R*(*t*) for the region where accepting population travel from other regions with higher *R*(*t*). In this scenario, we specify the *P* matrix to be:

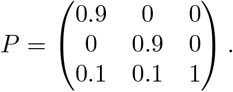

we specify the *R*(*t*) for the three regions to be (shown in Fig 1):

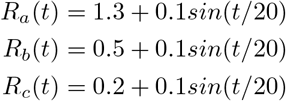

In this scenario, we initiate the 100 incidence for each of the three regions. We use Approach I to perform the estimation, since when the incidence counts are high, both approaches generate similar result. The result of estimated *R*(*t*) is shown in Fig S3 in S1 Appendix.

### Real Data Application

We implement the two approaches described above to the COVID-19 incidence data from the CDC. Since case reporting was more regular starting around July 2020, we focus on the case report data from July 2020 to March 2021. We aim to estimate the heterogeneous instantaneous reproductive numbers for all counties (14 in total) in Massachusetts, USA.

Human mobility patterns across the counties are examined, followed by the estimation of instantaneous reproductive numbers as well as the expected incidence for each county. While performing the estimation, we assume the serial interval follows a gamma distribution Gamma(3.45, 0.66), which corresponds to a mean of 5.2 days and a SD of 2.8 days [19].

## Results

Our approaches are based on the renewal equation framework proposed by Fraser et al. [2] for estimating the instantaneous reproductive number. We extend the framework to incorporate human mobility data in a system of renewal equations to estimate the instantaneous reproductive numbers for multiple regions. We propose two approaches to carry out the estimation. For Approach I, we adjust the incidence in multiple regions according to the human mobility data and then estimate the instantaneous reproductive number separately in each region using the EpiEstim method. We call this the incidence adjustment approach. For Approach II, we perform estimation using a system of renewal equations in a hierarchical Bayesian framework. We call this the Bayesian approach.

In this section, we show results for both the simple incidence adjustment approach and the more complex system of renewal equations using simulation and data from Massachusetts during the COVID-19 pandemic.

### Simulation Results

Our simulation study considers three regions with substantially different transmission profiles over time, but reasonably similar patterns in incidence. The incidence data are simulated with pre-specified reproductive numbers over time as well as a transition matrix, which can be informed by mobility data in practice, that describes how the population in each region distribute to other regions. The details of the simulations are described in Section. Approach I is straightforward as we use the human mobility data deterministically to adjust the incidence. For Approach II, we evaluated the model using different assumptions on the distribution of incidence and randomness for the mobility data. In this section, we show the results from the two proposed approaches along with the naive approach that does not use mobility data in Fig 2. The naive approach estimates the reproductive numbers separately for each region, which is equivalent to Approach I and Approach II without using the mobility data. Other simulation results are in the appendix.

**Fig 2.**
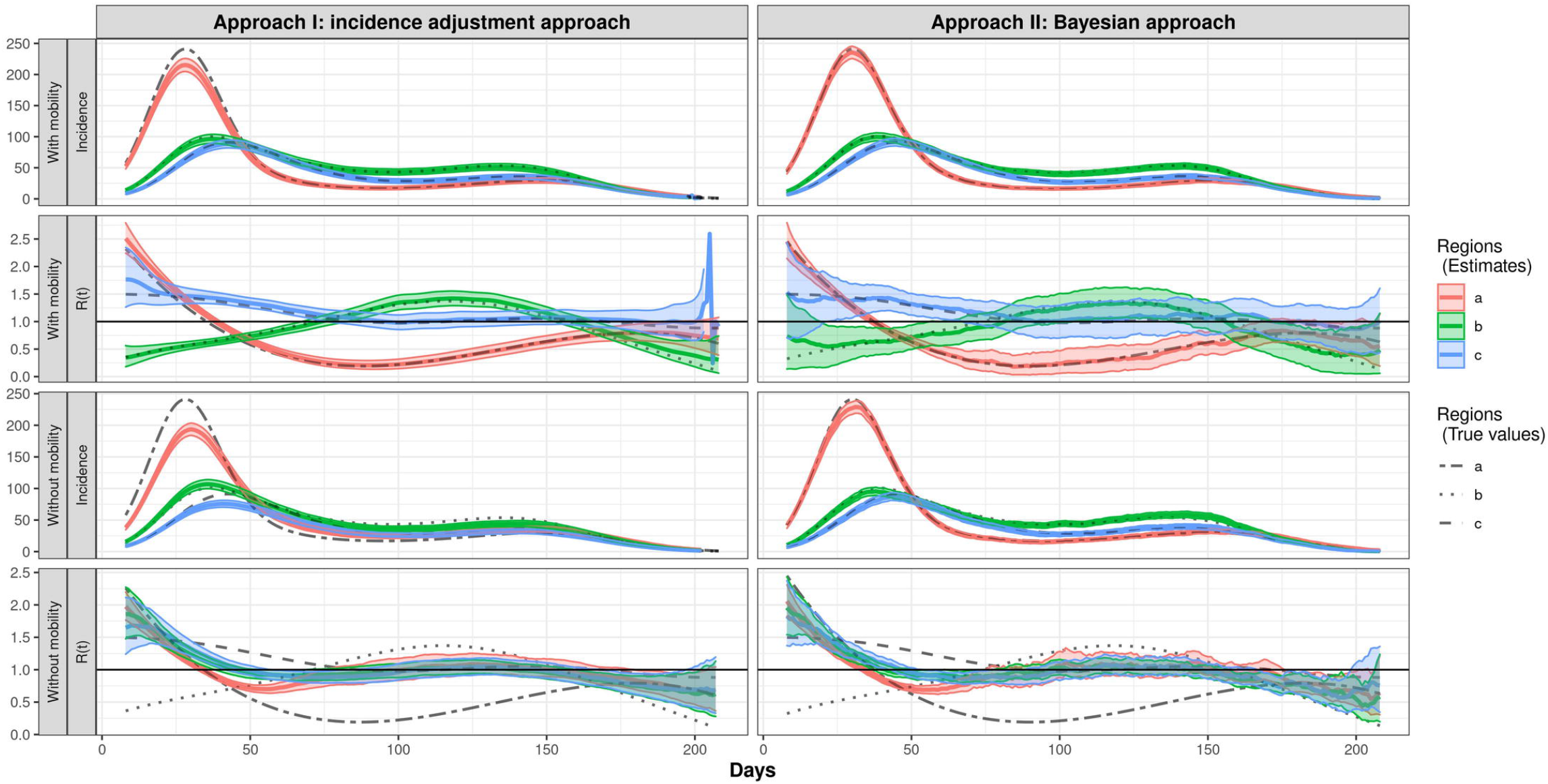
Estimated Incidence and *R*(*t*) by region. Solid lines are posterior means, along with the 95% credible bands (shaded).

Fig 2 shows the main result of the simulation. For the incidence adjustment approach, a fixed transition matrix *P* (for mobility between regions) is used for the estimation, and for the Bayesian approach, Dirichlet priors with concentration parameter 10^4^ are placed on the row vectors in the transition matrix *P*. Both of the methods provide estimated incidence for the 3 regions that are close to the incidence mean for 100 Monte Carlo replicates. The estimated reproductive numbers are also close to the true reproductive numbers. The credible bands of the estimated reproductive numbers are quite narrow for the incidence adjustment approach, while it is wider in the Bayesian approach.

When we do not use mobility data (i.e. *P* is an identity matrix), the incidence estimates obtained by Approach I deviate from the true incidence mean, especially earlier in the outbreak, compared to that obtained by Approach II. Although the estimated incidences obtained by Approach II are close to the mean of simulated data, the *R*(*t*) estimates obtained by both approaches when not accounting for mobility, are very similar for each of the three regions and quite different from the true *R*(*t*) curves. The results show that the estimates for *R*(*t*) are close to the true *R*(*t*) if we use the mobility information in the model. But if we just stratify the data by region and estimate *R*(*t*) ignoring mobility patterns between regions, we are not able to capture the transmission differences.

### COVID-19 Results

#### Overview of incidence in Massachusetts

Fig 3 is an overview of the incidence of COVID-19 from July 1^st^, 2020 to Feburary 28^th^, 2021 for the 14 counties in the State of Massachusetts, USA. Essex, Middlesex and Suffolk county, the most populous counties, have relatively high incidence. The overall pattern of incidence across counties is similar, exhibiting an obvious increase after November 2020 during the second wave.

**Fig 3.**
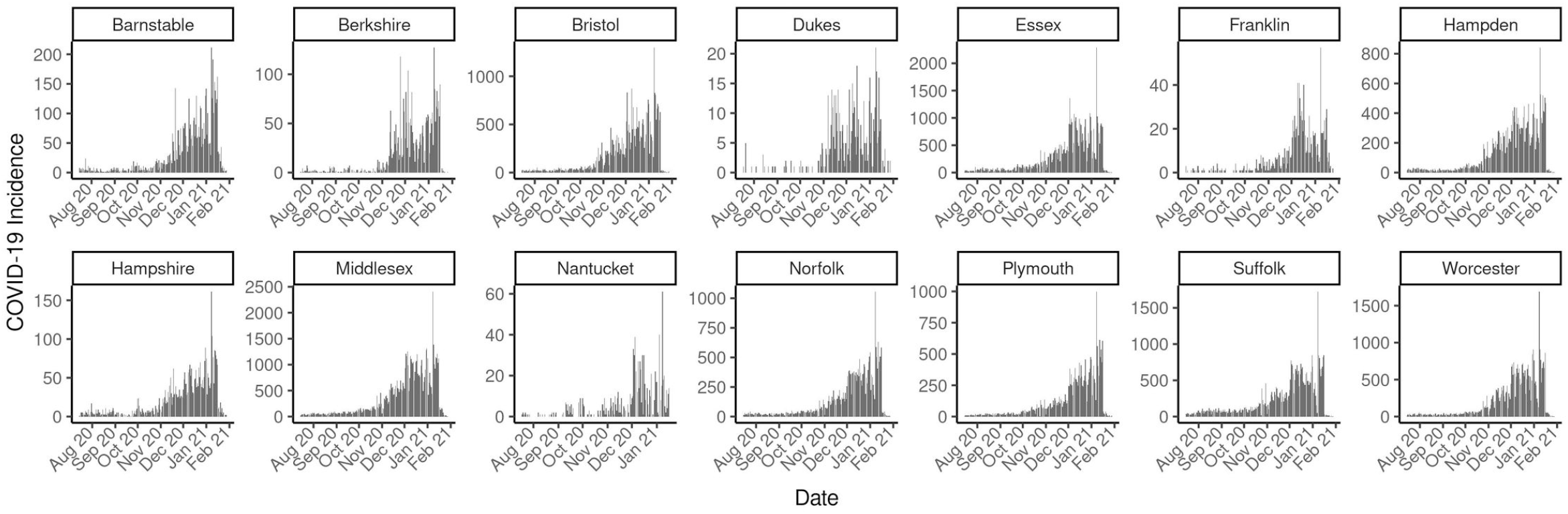
Reported Incidence for all MA counties.

#### Population flow across counties in Massachusetts based on human mobility data

Human mobility data is obtained from the multiscale dynamic human mobility flow dataset constructed and maintained by Kang et al. [16]. They computed, aggregated and inferred the daily and weekly dynamic origin-to-destination (O-D) flow at three geographic scales (census tract, county and state) analysing anonymous mobile phone users’ visits to various places provided by SafeGraph [17]. We use county level data in Massachusetts for the modeling in this real data analysis. The human mobility data consists of the estimated number of visitors traveling from one county to another in each day. We use *L*_*ij*_(*t*) to denote the number of visitors from county *j* to county *i* in day *t*. To visualize how the counties are clustered according to visitors traveling between them, we compute the average daily population flow 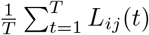 for each county pair from July 1^st^, 2020 to February 28^th^, and stratify the flow by weekdays and weekends, assuming there will be different patterns for working days and non-working days.

There are notable differences between weekday and weekend patterns of mobility that can be seen in the heatmaps and dendrograms (Fig 4) generated with complete-linkage hierarchical clustering. During weekdays, most travel is between regions that are geographically proximate, for example Barnstable, Bristol and Plymouth. On weekends, counties further apart are in the same cluster on the heat map, for example, Norfolk is in the cluster with Barnstable, Bristol and Plymouth. We also show the population flow on the geographical map in Fig 5. The clustering is more clear for regions that are geographically proximate for the daily average that is not stratified by weekdays and weekends. From the figure showing the difference between the weekdays’ daily average and weekends’ daily average, we observe that there is more of the population traveling between Essex, Worcester, Norfolk, Sulfolk and Middlesex on weekdays compared to weekends, and less of the population traveling between Middlesex, Barnstale and Plymouth as well as between Norfolk, Barnstale and Plymouth. These patterns support the clustering in the heat maps.

**Fig 4.**
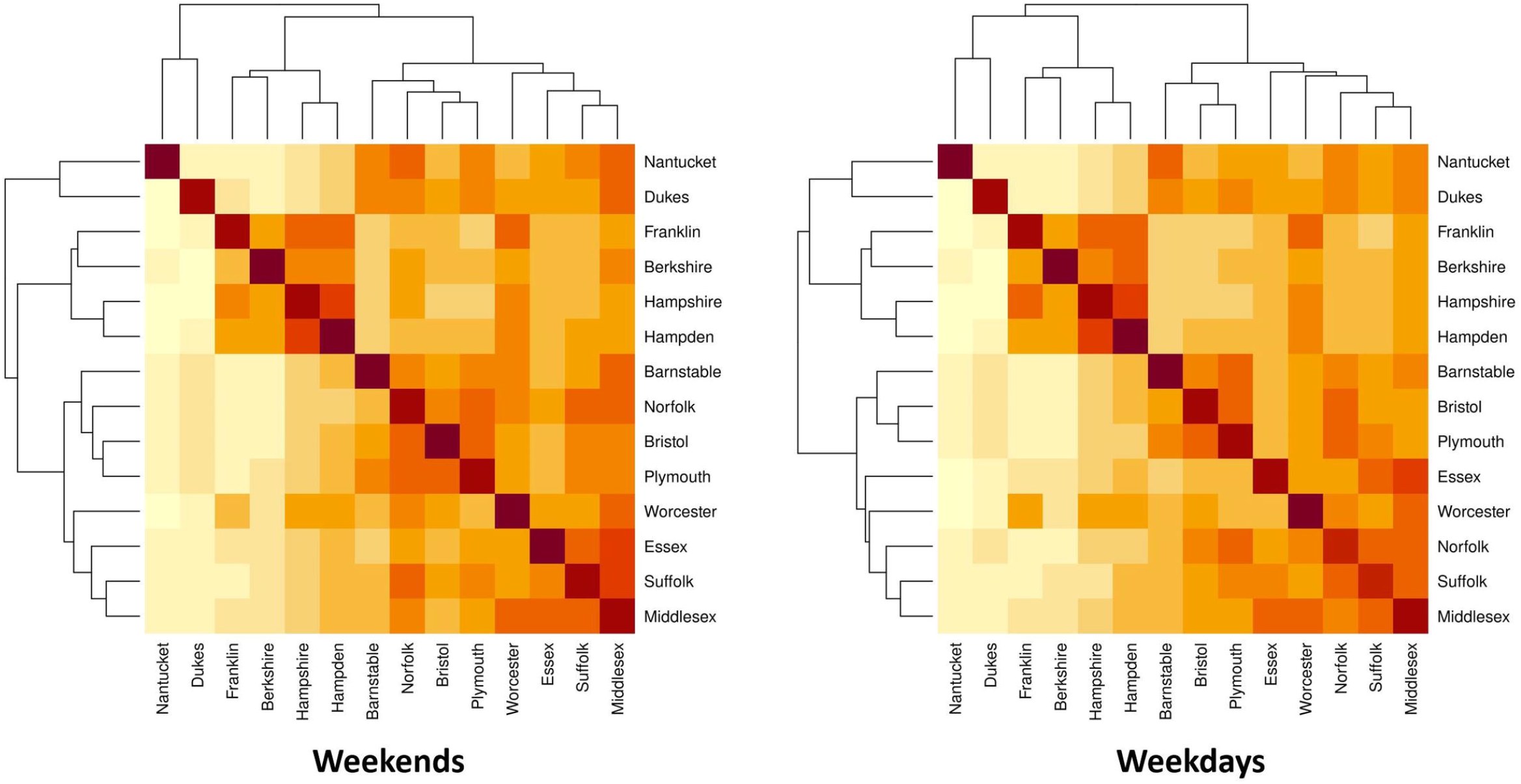
Heat maps for average population flow (log scaled) across regions during weekdays and weekends. Darker colors indicate regions with more flow between them.

**Fig 5.**
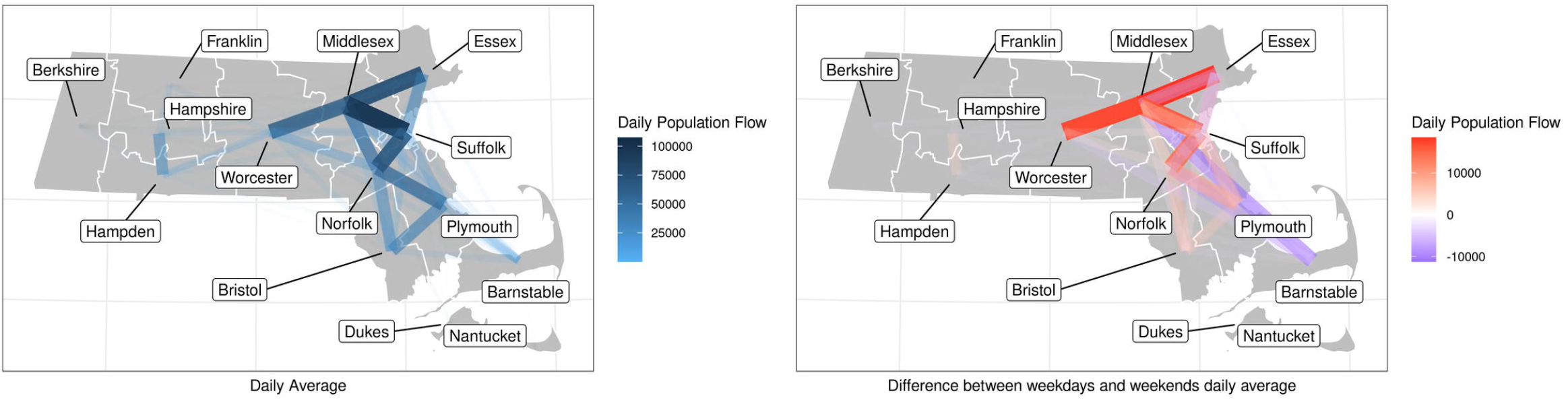
Human mobility network among counties of Massachusetts. The figure on the left shows the average daily population flow, and the figure on the right shows the difference of average population flow between weekdays and weekends, a positive value means the population flow is larger for weekdays than weekends, and a negative value means the population flow is smaller for weekdays than weekends

To quantify the daily change of population, for day *t*, we can aggregate the number of visitors from one county that travel to other counties to be the size of population flow out in day *t*, and denote it as *L*_out_(*t*). Also, we can aggregate the number of visitors from other counties that travel to that county to be the size of population flow in on day *t*, and denote this as *L*_in_(*t*). And we use *L*(*t*) to denote the population size on day *t*. Note that the data is for the human mobility on that day, instead of a permanent move.

Population change can inform how the population in a region is mixing with other regions, so we examine the percentage of population change for all counties as shown in Fig 6. Population change refers to the change in population size in day *t* compared to the population size in day *t* − 1 as a ratio, that is 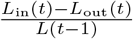, where *L*_in_ denotes the size of population flow in and *L*_out_ the size of population flow out. The population change plot shows that there is a relatively high percentage of population change for Barnstable, Dukes and Nantucket before October, 2020, due to the population inflow.

**Fig 6.**
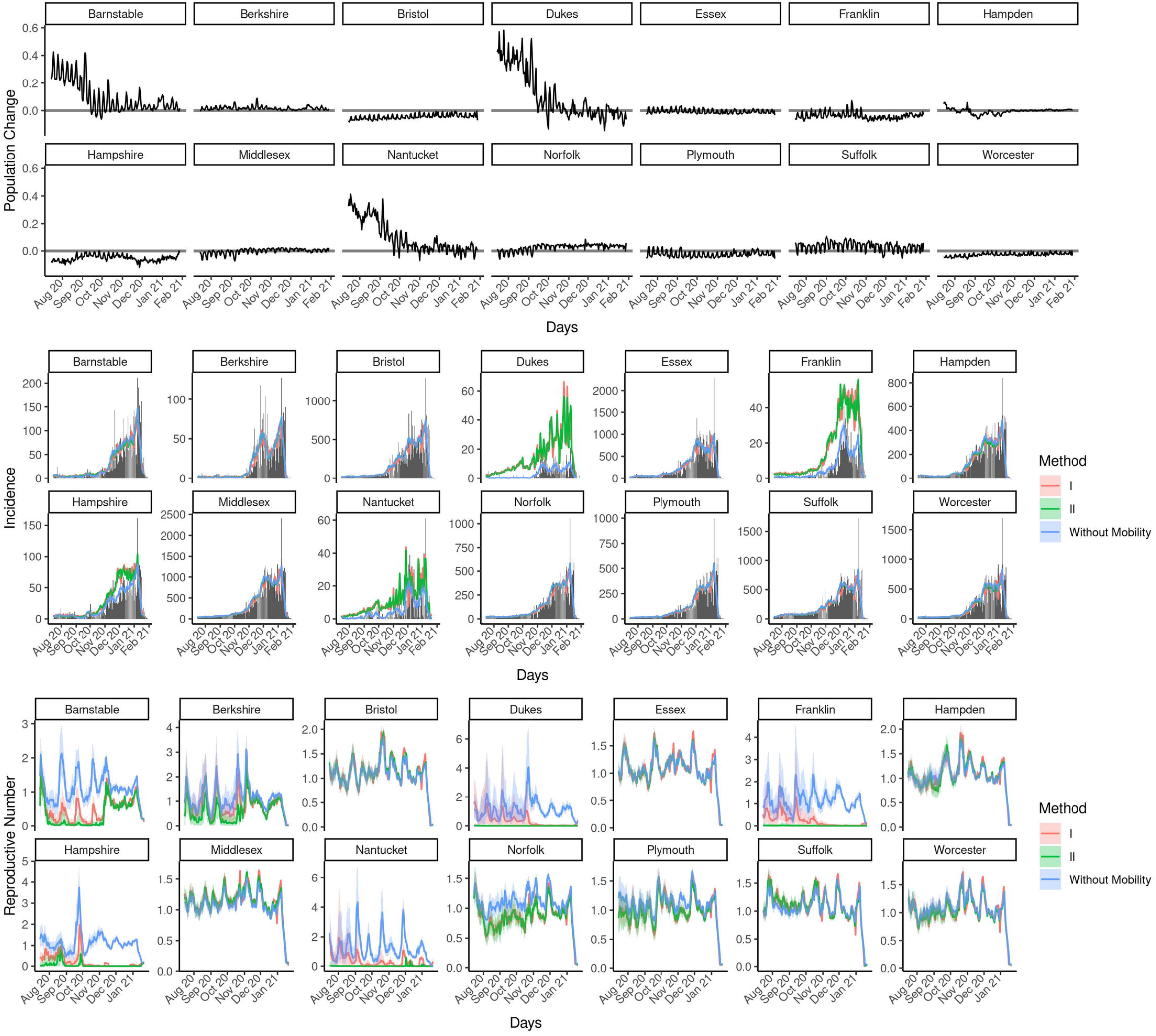
Population change, estimated Incidence and *R*(*t*) for all MA counties. Solid lines are the posterior means for incidence and *R*(*t*), along with the 95% credible band. The bar plots for the observed incidence are also shown. Results from Approach I, the incidence adjustment approach, are in red and those from Approach II, the Bayesian approach, are in green, and those from the original Fraser’s method (obtained by Approach II without incorporating mobility data) are in blue.

#### Estimated expected incidence and heterogeneous instantaneous reproductive numbers

Fig 6 shows the estimated expected incidence and *R*(*t*) for all counties with both of our proposed methods. Results from Approach I, the incidence adjustment approach, are shown in red and those from Approach II, the Bayesian approach, are shown in green, and the results from original Fraser’s approach (obtained by Approach II without incorporating mobility data) are shown in blue. The estimated *R*(*t*) are relatively lower for Barnstable, Dukes, Franklin, Hampshire and Nantucket. Three of these counties (Barnstable, Dukes and Nantucket) have a larger percentage of population flow in from July to October. From the result, it is possible that the increase of incidence in these counties is due to inflow from counties with higher *R*(*t*). The results from Approach II are similar to those from Approach I when R(t) is high, while in counties with lower reproductive numbers the results from Approach II are even smaller than Approach I.

## Discussion

It is well-established for many infectious diseases that there is substantial heterogeneity in transmission patterns. One might reasonably expect that some of this variability occurs geographically due to a potentially complex combination of social factors and some amount of stochastic effects. Estimating spatially granular reproductive numbers allows for greater targeting of interventions and the potential to uncover the factors that drive heightened transmission. We have described two approaches for estimating *R*(*t*) that incorporate mixing patterns between distinct groups, which in our setting is informed by mobility data between geographic regions.

We demonstrate how these two approaches perform on simulated data. Simulations shows that both of the approaches are able to estimate the heterogeneous instantaneous reproductive numbers for multiple regions well when the mobility data is well-specified. We observed that the second approach has larger variability. This is expected since in Approach II we incorporate some of our uncertainty around the accuracy of the mobility data, allowing some flexibility in the case where the mobility data might not exactly represent how incident cases are flowing between the regions. This means that the first approach is likely more sensitive to inaccuracies in the mobility data, while the second approach samples over for the mobility prior together with the other parameters, allowing for some misspecification. Therefore, if we have high quality mobility data that is representative of the population and incidence flow, and are only interested in obtaining reproductive numbers for multiple regions, we can use the more efficient Approach I. If we want to incorporate uncertainty in the mobility data and/or investigate factors that are associated with *R*(*t*), we can use Approach II.

In our simulation, we show that using mobility information allows us to obtain estimates for *R*(*t*) that are close to the true *R*(*t*) and that this is not feasible when mobility data is not used (see scenario 1 in supplement). In other words, simply stratifying data by region and estimating *R*(*t*) ignoring mobility patterns between regions does not appropriately capture transmission differences. This is especially important when there are regions with a lower *R*(*t*) accepting population flow from regions with a higher *R*(*t*). For example, people might live in counties with lower *R*(*t*), but work in counties with higher *R*(*t*). If mobility information is not taken into account, we could over estimate *R*(*t*) for the counties in which these people are living, and under estimate the *R*(*t*) in the counties where they work. This is shown in our simulation results (see scenario 3 in supplement). We realize that there could be low incidence counts for some regions, and we showed that the Approach I tends to overestimate *R*(*t*) compared to Approach II in this setting (see scenario 2 in supplement). Both of the approaches have a larger credible band, which indicates the these approaches are not ideal in the presence of low incidence count.

A potential additional benefit of the more computationally intensive second approach is that local factors, such as age, social economic status and disease containment policies can be incorporated into the estimation framework. This can potentially allow one to not only estimate more accurately the differences between regions, but also potentially start to more carefully understand some of the underlying factors influencing the transmission differences.

When we consider the dynamics of COVID-19 in Massachusetts, the county-level results show that the two approaches yield similar estimates, but that these are distinct from the naive approach that ignores mobility. Generally, the estimated incidence data is similar, but there are some differences in the estimated *R*(*t*) with mobility incorporated. *R*(*t*) estimates from Approach I have larger credible band for the counties with lower incidence, such as Berkshire, Dukes, Franklin and Nantucket. The second approach produces smoother estimated *R*(*t*) when *R*(*t*) is small. From the simulation scenario 2, we have shown that the Approach I tends to overestimate *R*(*t*) compared to Approach II when there are low counts for incidence. This could be the reason for the larger *R*(*t*) estimates from Approach I for Berkshire, Dukes, Franklin and Nantucket during the time with low incidence count. However, for Berkshire, Dukes and Nantucket, we observe a positive population change from July 1^st^, 2020 to October 20^th^, 2020, so we are more confident in the result from Approach II, since it is more likely that the incidence in these counties are due to the population input from other counties with higher *R*(*t*).

For both of the methods, an important assumption is that the mobility data describes the flow of infectious individuals, even though it is not explicitly measuring this. This might not hold if individuals dramatically change their behavior when they are infectious. A potential approach to cope with this problem might be adding parameters informed by behavioral data among infectious individuals as weights to the mobility data to account for changed mobility due to the disease.

In a summary, the instantaneous *R*(*t*) is an important metric for infectious disease surveillance, since it provides a real-time description of the transmission dynamics among the population. While estimating *R*(*t*) for multiple regions, we expect to see heterogeneity. However, estimating the heterogeneity can be challenging when there is extensive population flow between regions leading to a mixing of the population that can mask or misrepresent the true transmission dynamics. We have presented two methods that incorporate mobility data for the estimation of spatially heterogeneous *R*(*t*). The ultimate goal of this approach is to identify the regions with higher transmission in which to focus interventions as well as study potential mechanisms of transmission. These methods have broad applicability to estimating *R*(*t*) in the presence of any potential heterogeneities, such as age-mixing which can use mixing behavior described by contact surveys such as those performed by Mossong et al. [20].

## Data Availability

The datasets generated for simulation and analyzed for real data analysis are available in the GitHub repository.

https://github.com/zwzhou-biostat/hrt/data

## Supporting information

**S1 Appendix. Other simulation results** Simulation result for Model 1, 2 and 3 in scenario 1, and results for scenario 2 and 3.

---

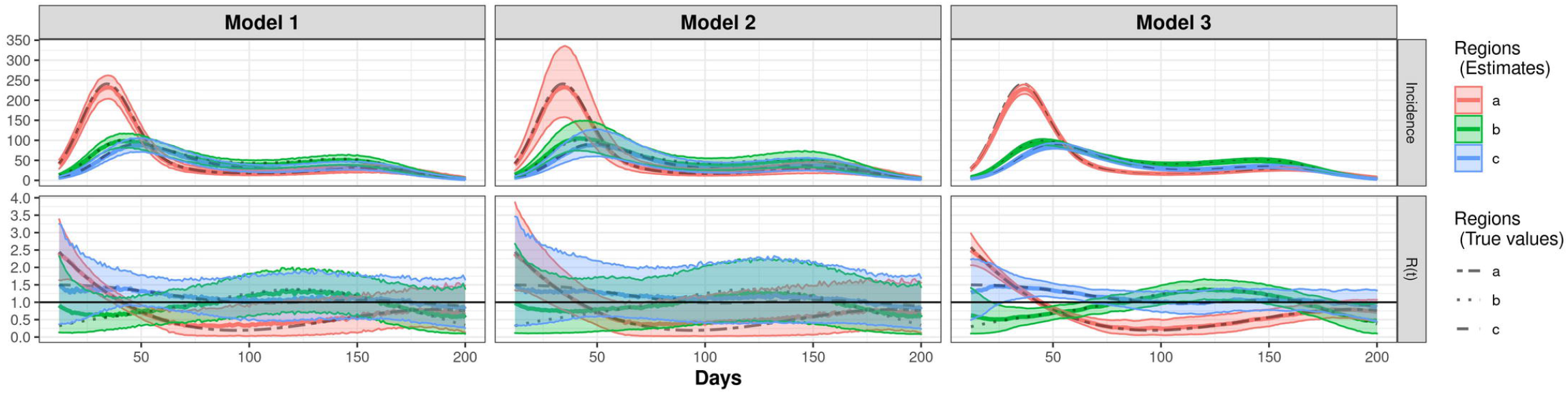

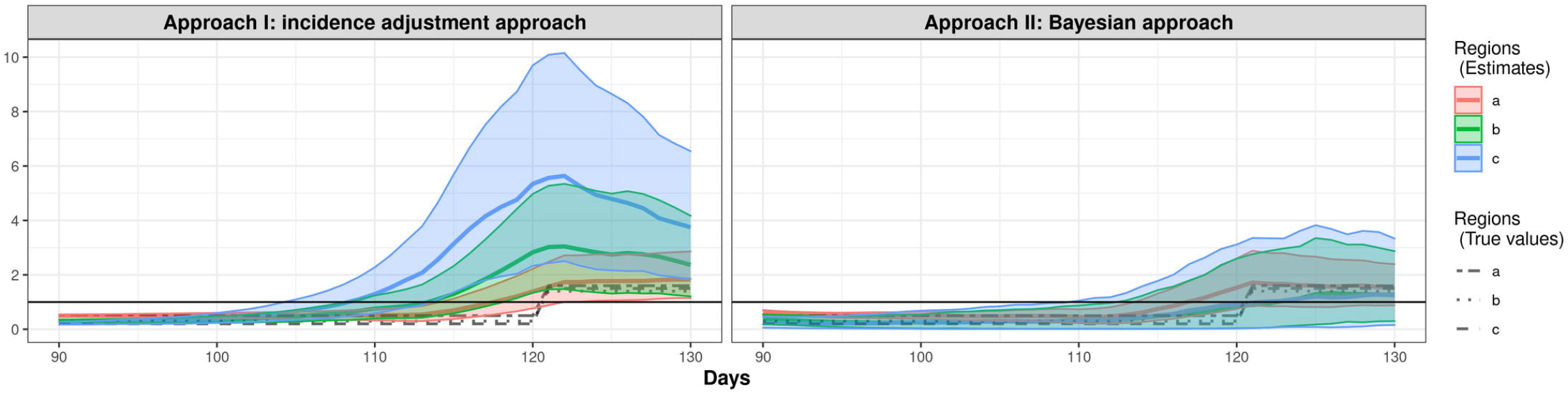

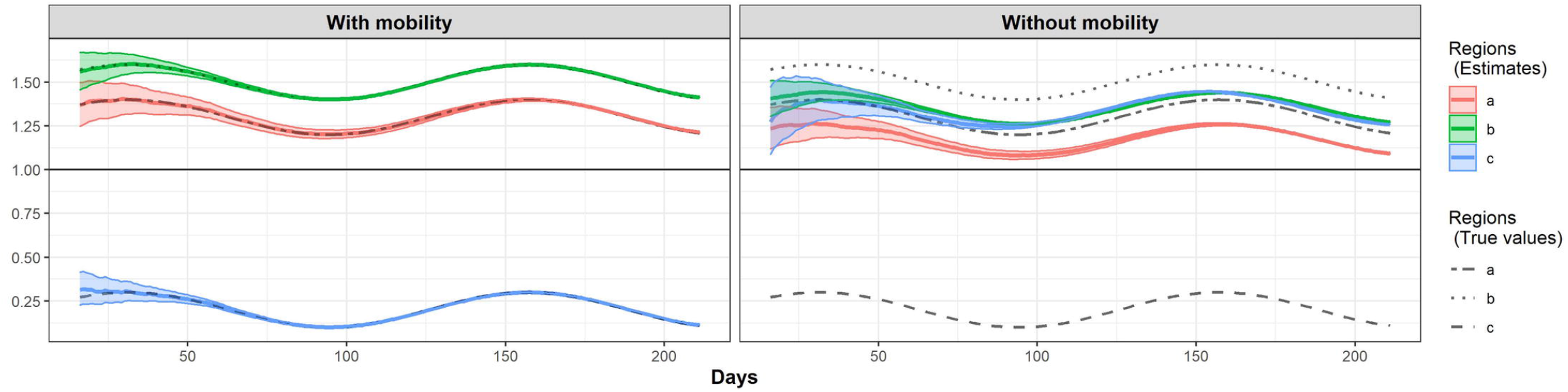

## References

1. Wallinga J, Teunis P. Different epidemic curves for severe acute respiratory syndrome reveal similar impacts of control measures. American Journal of epidemiology. 2004;160(6):509–516.

2. Fraser C. Estimating individual and household reproduction numbers in an emerging epidemic. PloS one. 2007;2(8):e758.

3. Cori A, Ferguson NM, Fraser C, Cauchemez S. A new framework and software to estimate time-varying reproduction numbers during epidemics. American journal of epidemiology. 2013;178(9):1505–1512.

4. Thompson R, Stockwin J, van Gaalen RD, Polonsky J, Kamvar Z, Demarsh P, et al. Improved inference of time-varying reproduction numbers during infectious disease outbreaks. Epidemics. 2019;29:100356.

5. Li T, White LF. Bayesian back-calculation and nowcasting for line list data during the COVID-19 pandemic. PLoS computational biology. 2021;17(7):e1009210.

6. Günther F, Bender A, Katz K, Küchenhoff H, Höhle M. Nowcasting the COVID-19 pandemic in Bavaria. Biometrical Journal. 2021;63(3):490–502.

7. Martinez de Salazar P, Lu F, Hay J, Gomez-Barroso D, Fernandez-Navarro P, Martinez E, et al. Near real-time surveillance of the SARS-CoV-2 epidemic with incomplete data. 2021;.

8. Pitzer VE, Chitwood M, Havumaki J, Menzies NA, Perniciaro S, Warren JL, et al. The impact of changes in diagnostic testing practices on estimates of COVID-19 transmission in the United States. MedRxiv. 2020;.

9. White LF, Archer B, Pagano M. Estimating the reproductive number in the presence of spatial heterogeneity of transmission patterns. International journal of health geographics. 2013;12(1):1–10.

10. Sun K, Wang W, Gao L, Wang Y, Luo K, Ren L, et al. Transmission heterogeneities, kinetics, and controllability of SARS-CoV-2. Science. 2021;371(6526).

11. Woolhouse ME, Dye C, Etard JF, Smith T, Charlwood J, Garnett G, et al. Heterogeneities in the transmission of infectious agents: implications for the design of control programs. Proceedings of the National Academy of Sciences. 1997;94(1):338–342.

12. Zhang Y, Li Y, Wang L, Li M, Zhou X. Evaluating transmission heterogeneity and super-spreading event of COVID-19 in a metropolis of China. International journal of environmental research and public health. 2020;17(10):3705.

13. Sy KTL, Martinez ME, Rader B, White LF. Socioeconomic Disparities in Subway Use and COVID-19 Outcomes in New York City. American Journal of Epidemiology. 2020;doi:10.1093/aje/kwaa277.

14. Mishra S, Kwong JC, Chan AK, Baral SD. Understanding heterogeneity to inform the public health response to COVID-19 in Canada. CMAJ. 2020;192(25):E684–E685.

15. CDC. Centers for Disease Control and Prevention, COVID-19 Response. COVID-19 Case Surveillance Data Access, Summary, and Limitations (version date: December 31, 2020); 2021. https://data.cdc.gov/Case-Surveillance/COVID-19-Case-Surveillance-Public-Use-Data/vbim-akqf.

16. Kang Y, Gao S, Liang Y, Li M, Kruse J. Multiscale Dynamic Human Mobility Flow Dataset in the U.S. during the COVID-19 Epidemic. Scientific Data. 2020; p. 1–13.

17. SafeGraph. The impact of coronavirus (COVID-19) on foot traffic; 2020. Available from: https://www.safegraph.com/dashboard/covid19-commercepatterns.

18. Mishra. On the derivation of the renewal equation from an age-dependent branching process: an epidemic modelling perspective. arXiv preprint arXiv:200616487. 2020;.

19. Zhang J, et al. Evolving epidemiology of novel coronavirus diseases 2019 and possible interruption of local transmission outside Hubei Province in China: a descriptive and modeling study. MedRxiv. 2020;.

20. Mossong J, Hens N, Jit M, Beutels P, Auranen K, Mikolajczyk R, et al. Social contacts and mixing patterns relevant to the spread of infectious diseases. PLoS medicine. 2008;5(3):e74.

