## Supplementary material for "Estimation of heterogeneous instantaneous reproduction numbers with application to characterize SARS-CoV-2 transmission in Massachusetts counties"

### Supporting information

#### Simulation result for Model 1, 2 and 3 in scenario 1

Fig S1 shows the estimated incidence and $R(t)$ by model 1, 2 and 3. In model 1, the incidences are assume to be following Poisson distribution. The estimated $R(t)$ have the trend that align with the true $R(t)$ curves simulated. And the predicted incidence for the 3 regions are close with the mean of incidence in the simulated datasets. Since there is no smoothing for the estimates in model 1, we also observe more variations of the estimates within a short period of time.

In model 2, we assume the incidences are following negative binomial distributions although the simulated data is from Poisson distribution. Credible band is wider than that in results from model 1 and the estimates are also not smooth as that in results from model 1.

In model 3, incidences are assumed to be following Poisson distribution. The trend of $R(t)$ estimates are also aligning with the true $R(t)$, and the estimates are smoother with a smoothing window of 8 days in the model. The credible band of the posterior estimates are narrower.

**Fig S1. Estimated Incidence and $R(t)$ by Model 1, 2 and 3.**

#### Simulation result for scenario 2: low incidence count

**Fig S2. Estimated $R(t)$ for low count scenario**

Fig S2 shows the estimated $R(t)$'s for the three regions by both Approach I and Approach II. Both of the approaches have wider credible band when the incidence counts are low from day 110 to day 130. For Approach I, the estimated $R(t)$ are much larger than the true $R(t)$ with low incidence counts, for example, for region a, the posterior mean of $R(t)$ is 5.8 and the true $R(t)$ is 1.6 at day 125. For Approach II, the estimated $R(t)$ is closer to the true $R(t)$, for example, for region a, the posterior mean of $R(t)$ is 1.72 and the true $R(t)$ is 1.6 at day 125.

### Simulation result for scenario 3: population input from other regions

**Fig S3. Estimated $R(t)$ for population input from other regions**

In the scenario where the population travel from two regions with higher $R(t)$ to the third region with a lower $R(t)$, we observed the difference for the estimated $R(t)$ when using Approach I with mobility information and without mobility information, note that Approach I without mobility information is equivalent to the original Fraser's method. There is an overestimate for the $R(t)$ in the region accepting population from other two regions.
